# “Hurts less, lasts longer” experiences of young people receiving high-dose subcutaneous infusions of benzathine penicillin G to prevent rheumatic heart disease

**DOI:** 10.1101/2023.09.13.23295467

**Authors:** Julie Cooper, Stephanie L Enkel, Dhevindri Moodley, Hazel Dobinson, Erik Andersen, Joseph H Kado, Renae K Barr, Sam Salman, Michael G Baker, Jonathan R Carapetis, Laurens Manning, Anneka Anderson, Julie Bennett

## Abstract

**Background:** Four-weekly intramuscular (IM) benzathine penicillin G (BPG) injections to prevent acute rheumatic fever (ARF) progression have remained unchanged since 1955. A Phase-I trial in healthy volunteers demonstrated the safety and tolerability of high-dose <u>S</u>ub<u>C</u>utaneous Infusions of B<u>P</u>G (SCIP) which resulted in a much longer effective penicillin exposure, and fewer injections. Here we describe the experiences of young people living with ARF participating in a Phase-II SCIP trial.

**Methodology:** Participants (n=20) attended a clinic in Wellington, New Zealand (NZ). After a physical examination, participants received 2% lignocaine followed by 13.8mL (6 vials) to 20.7mL (9 vials) of BPG (Bicillin-LA^®^; determined by weight), into the abdominal subcutaneous tissue. Semi-structured interviews and observations were taken during and after the infusion, as well as on days 28 and 70. All interviews were recorded, transcribed verbatim, and thematically analysed.

**Principal Findings:** Low levels of pain were reported on needle insertion, during and following the infusion. Some participants experienced discomfort and bruising on days one and two post dose; however, the pain was reported to be less severe than their usual IM BPG. Participants were ‘relieved’ to only need injections quarterly and the overwhelming majority preferred to continue with SCIP.

**Conclusions:** Participants preferred SCIP over their usual regimen, reporting less pain and a preference for the longer time gap between treatments. Recommending SCIP as standard of care for most patients needing long-term ARF/RHD prophylaxis has the potential to transform secondary prophylaxis of ARF/RHD in NZ and globally.

**Synopsis:** Acute rheumatic fever (ARF) is a preventable inflammatory disease that occurs as a delayed sequelae to group A streptococcus (GAS) infection. ARF and its complication rheumatic heart disease (RHD) have significant negative effects on health, often resulting in chronic illness and premature death. For 70 years, the only proven way to prevent ARF progression has been benzathine penicillin G (BPG), given as a monthly intramuscular (IM) injection for a minimum of 10 years. The effectiveness of this approach is limited by pain and the frequency of injection which leads to suboptimal adherence. There is an urgent need to improve penicillin formulations for all children living with ARF and RHD. Here we describe the experiences of 20 young people living with ARF participating in a Phase-II trial delivering high-dose <u>S</u>ub<u>C</u>utaneous Infusions of <u>P</u>enicillin (SCIP) in order to provide longer effective penicillin exposure, and therefore fewer injections. Participants in the trial overwhelmingly preferred high-dose SCIP over their usual monthly IM penicillin regimen, reporting less pain and a preference for the longer time gap (28 versus 70 days) between treatments. Reducing injection frequency from 13 to four-or-five per year, may improve adherence and reduce disease progression. Offering widespread SCIP to ARF/RHD patients to evaluate long-term adherence, preferences and disease progression has the potential to transform secondary prophylaxis of ARF/RHD both in New Zealand and globally.

## Introduction

Acute rheumatic fever (ARF) is an immune-mediated inflammatory disease that occurs as a delayed sequela to group A streptococcus (GAS) infection [1, 2]. A single acute or several episodes of ARF can progress to rheumatic heart disease (RHD); a serious condition characterised by permanent heart valve damage that may result in early death. It is estimated that 33.4 million people worldwide live with RHD, resulting in approximately 319,400 deaths each year [3]. ARF and RHD have all but disappeared from high-income countries, yet in Aotearoa New Zealand (NZ), they remain an alarming and inequitable cause of preventable suffering and death for Indigenous Māori and Pacific Peoples [4].

Since the 1950s, benzathine penicillin G (BPG) has been used extensively for the treatment of several infectious diseases - including ARF and RHD – with a unique and useful characteristic being its prolonged serum concentration. Referred to as secondary prophylaxis [5, 6], four-weekly intramuscular (IM) injections of BPG [7] are needed to ensure plasma penicillin concentrations remain above 0.02mg/L (20ng/mL), a pharmacological surrogate of protection against repeated GAS infections that may worsen disease [8]. Current NZ guidelines recommend patients with ARF have a minimum 10 years of secondary prophylaxis or until the patient is aged 21 years for mild disease, 30 years for moderate or 40 years for severe cases [9]. It is also recommend that patients receive at least 80% of secondary prophylaxis injections (11 of 13 each year); however, reported adherence is much lower than this [10].

The pain associated with IM BPG injection is frequently cited as a reason for lack of adherence to secondary prophylaxis [11, 12]. Therefore, the addition of lignocaine to reduce injection pain has been recommended by the NZ Ministry of Health [13]. Additionally, application of pressure and temperature packs are two methods that have shown significant reduction in pain scores with IM BPG injections [11, 14, 15]. However, ultimately to improve adherence and prevent disease progression there is an urgent need to improve the delivery and formulation of long acting penicillins [16].

A consultation with global experts in RHD suggested changing formulations of BPG to produce a product that is longer acting, less painful, and/or more reliable in its pharmacokinetics [12, 17]. They concluded that an acceptable reformulation would need to: be administered subcutaneously; have a dosing schedule greater than six weeks; be less or no more painful than existing BPG; be cold-chain independent; and of comparable cost to IM BPG [12].

Recent work has found subcutaneous (SC) delivery of BPG to be safe and potentially advantageous. In a randomised cross-over trial, Kado et al., [18] compared the pharmacokinetic profile and tolerability of BPG delivered by IM and SC routes of administration. Subcutaneous delivery was superior, having a more prolonged duration of effect, comparable pain scores and no adverse effects [18].

Building on this trial, it was predicted that SC infusion of high-dose BPG could provide adequate penicillin concentrations for up to three months. To test the safety and tolerability of high-dose <u>SC</u> Infusion of benzathine <u>P</u>enicillin (SCIP-I), a dose escalation Phase-I trial was conducted in 24 healthy adults [19]. The study concluded that delivering high-dose SCIP was safe, had acceptable tolerability and could be suitable for up to three-monthly dosing intervals for secondary prophylaxis of ARF/RHD [19]. In addition to the Phase-I trial, a qualitative sub-study was undertaken to provide in-depth information about the tolerability and acceptability of SCIP [20]. The sub-study demonstrated that SCIP was acceptable to participants, and while some experienced higher pain levels, most had tolerable mild discomfort. Potential approaches to alleviate pain and discomfort were explored and suggestions included distractions, a slower infusion time and larger doses of lignocaine anaesthesia.

Incorporating the learnings from the SCIP-I, progression to a Phase-II study investigating the delivery of high-dose SC BPG was undertaken in NZ children and young adults with ARF currently receiving secondary prophylaxis (SCIP-II). This paper reports the findings of a qualitative SCIP-II sub-study aiming to assess the acceptability of SCIP compared to the current regimen experienced by participants.

## Methods

The methods of the SCIP-I trial have been described elsewhere[19] with SCIP-II following a similar approach. In brief, 20 participants with ARF and prescribed to receive four-weekly IM BPG attended an outpatient clinic in Wellington, NZ. Following a physical examination, participants received 2% lignocaine (up to 5mLs) to the subcutaneous abdominal space through a 22G Saf-T-Intima Cannula. This was followed by 13.8mL (7.2 million units [MU]; 6 vials) to 20.7mL (10.8 MU; 9 vials) of BPG delivered via a series of slow manual pushes, from manufacturer’s prefilled 2.3mL glass syringes (Bicillin-LA^®^, Pfizer) [21], dosed according to participant’s weight (Table 1). One difference in the BPG administration between SCIP-I and SCIP-II was that to deliver Bicillin-LA^®^ SCIP-I used a spring-driven syringe infusion pump (Springfusor 30, Go Medical Industries Pty Ltd., Subiaco, Australia) with the use of a variable flow control device (VersaRate Plus, EMED Technologies, El Dorado Hills, California, USA). SCIP-II opted instead to use a series of slow, steady pushes, as this enabled better control over the infusion speed and mitigated the need to transfer the contents of the pre-filled syringes into a larger syringe.

**Table 1.** Bicillin-LA dosing for subcutaneous infusion.

| Weight (kg) | Number of vials | Volume (mL) |
| --- | --- | --- |
| 30-50 | 6 | 13.8 |
| 50-70 | 7 | 16.1 |
| 70-90 | 8 | 18.4 |
| >90 | 9 | 20.7 |

### Data collection

The study applied a qualitative Kaupapa Māori research design and included individual semi-structured interviews and participant observations. Kaupapa Māori research is a critical framework that gives meaning to the life of Māori and analyses unequal relations of power that influence Māori wellbeing. Such a framework allows for an empowering lens that places Māori at the centre of the study and rejects cultural deficit explanations [22, 23].

Participant observations were undertaken alongside audio-recorded participants interviews, which were conducted at three-time points; day 0 (during and after the infusion), day 28 and day 70 following dosing. In addition, participant observations were recorded on days one and two and demographic data were collected via case report forms. All interviews occurred face-to-face, with the first taking place at the bedside in the outpatient department, and the last two in the community (generally the participant’s home). Observational data were collected in a field journal by the study nurse or researcher, transcribed and analysed as described below. The semi-structured interviews used a standardised interview guide consisting of a series of open-ended questions regarding experience of the infusion, tolerability of the procedure, pain during and following the infusion, and comparisons to their usual IM BPG injections. To supplement the semi-structured interviews, individual audio-recorded informal interviews were had with the study nurse, a community nurse, and two researchers.

Quantitative pain was measured using a numerical rating score (NRS) on a 0-10 scale during participant interviews: 0 and 10 represented ‘no pain’ and ‘worst pain imaginable’, respectively. We considered minimal clinically-important differences (MCID) for moderate pain (NRS 4-7) and severe pain (NRS 8-10) to be 1.3 and 1.8, respectively as reported, although there is no accepted MCID for mild pain (NRS 1-3) [24].

### Data analysis

Interviews were transcribed verbatim along with field observations. Pseudonyms are used in place of names. Transcripts and observations were read repeatedly by one investigator who conducted the majority of the interviews and highlighted initial codes using NVIVO 12 [25]. Thematic saturation was defined as when new incoming data produced little to no new information on the topic under exploration. For some themes saturation was reached with the first 11 participants and no further themes were identified after 15 interviews. Included in the general inductive data analysis, were observations and experiences captured from those present during the infusions, including those from the study nurse, a community nurse, and two researchers. Raw qualitative data codes and themes were presented to Māori and Pacific researchers to assess cultural meaningfulness.

### Ethical approval

The ethical considerations of the study were approved by the NZ Health and Disability Ethics Committee (11094) and endorsed by Te Whatu Ora, Capital Coast and Hutt Valley Health, which included review by their Māori Research Board (RAG-M #916). In addition, the trial is registered with the Australian New Zealand Clinical Trials Registry (ACTRN12622000916741). All participants (or their parent / legal guardian) provided written, informed consent prior to participating, with information and consent forms available in Te Reo Māori and Samoan.

## Results

### Characteristics of participants

Of the 20 participants, the mean age was 15 years (range 6-32 years). Most participants were female (14, 70%). The majority (12, 60%) identified as Pacific (9 Samoan, 3 Tongan), with seven Indigenous Māori (35%), and one NZ European (Table 2). On average the infusion took 15 minutes (range 9-25 minutes).

**Table 2.** Characteristics of research participants.

| Pseudonym | Sex | Age band (years) | Ethnicity |
| --- | --- | --- | --- |
| Leilani | Female | 20-24 | Samoan |
| Anaru | Male | 15-19 | Māori/NZ European |
| Matisse | Female | 15-19 | Samoan |
| Sana | Male | 5-9 | Samoan |
| Mika | Male | 15-19 | Samoan |
| Mia | Female | 10-14 | Samoan |
| Hana | Female | 20-24 | Māori |
| Ani | Female | 15-19 | Māori |
| Talia | Female | 20-24 | Samoan |
| Lucy | Female | 15-19 | NZ European |
| Mele | Male | 10-14 | Tongan |
| Aroha | Female | 15-19 | Māori |
| Sione | Male | 10-14 | Tongan |
| Lulu | Female | 10-14 | Samoan |
| Marino | Male | 10-14 | Māori/NZ European |
| Nina | Female | >24 | Māori |
| Sefina | Female | 10-14 | Samoan |
| Natia | Female | 10-14 | Samoan |
| Kiri | Female | 10-14 | Māori |
| Langi | Female | 10-14 | Tongan |

### Thematic analysis

The thematic analysis revealed that although there was some initial anxiety from participants, in general, they experienced less pain with SCIP delivery when compared to IM. There was a strong preference to remain on SCIP. These findings are further described across six themes below.

### Anxiousness prior to SCIP

Anxiety was evident for many participants. As most of the participants had become used to having their IM BPG injections, coming in for an unknown procedure was an uncomfortable experience, with many participants arriving anxious, as described by Talia and Leilani:

> *“A bit nerve wracking. I was a bit anxious. But it was okay, there was no pain.”*
>
> *“I thought it would hurt more; I was overthinking it.”*

Developing a rapport with SCIP providers early on was key to reducing participant’s anxiety. Likewise, offering a koha (acknowledgement) in recognition of participant contribution and expenses (travel, parking), plenty of kai (food), iPads and Wi-Fi as distractions, and referral of whānau (families) to support services when needed, were all reported by participants as key facilitators of rapport and anxiety reduction. As the study progressed the Research Nurse was also able to reassure participants about some of the positive experiences from the early participants.

### Little pain during the needle insertion

For most participants inserting the needle containing the analgesia did not cause a lot of pain, it was often noted that ‘I barely felt it’ or described it as a tickle/tingle. For participants who did feel pain, it was described as a short, sharp, or stingy sensation that quickly subsided. Sana and Leilani describe their experiences:

> *“I barely felt it, didn’t hurt, bit of a pinch” [from nurse holding skin]*.
>
> *“Quite sharp pain didn’t last very long, just when it goes into the skin then can’t feel it after that.”*

### Little or no pain during the subcutaneous infusion

The majority of participants described having no or very little pain during the infusion. This was validated through the NRS where half of participants reported no pain at all during or after the infusion (NRS of 0). It was noted by the nurse and the researchers that many of the participants relaxed once the infusion started and many participants became sleepy during the procedure. Hana, Sana and Matisse describe that the infusion was not painful:

> *“No pain at all, just felt nurses’ hand on my tummy, [when needle went in]. During the process I didn’t notice it at all to be honest, I could see it but couldn’t feel it.”*
>
> *“No pain” (smiling)*
>
> *“Don’t feel anything, no pain”*

However, for some participants the infusion was associated with pain, which was often described as stinging, hard or numb. The pain was validated through pain scores which ranged from NRS 1-5. Sione described the infusion as:

> *“Weird, hot as it was going in. Burning, stinging. It stops and goes. Halfway through pain went up then when finished it went down.”*

For a small minority the pain increased during the infusion from low levels to sometimes quite painful nearing the end. Near the end of the infusion Nina described feeling:

> *“A little bit uncomfortable now, it is like a pull pain, like the volume expanding.”*

Two of the participants were in a state of discomfort with pain scores of seven. Without an understanding of participants’ pain tolerance, it was hard to know how much of the pain was a result of anxiety to the procedure. By the time they went home they were all feeling more comfortable (pain scores all below three). Aroha explains how she was feeling.

> *“It is so sore going in, stingy sore like a bee sting. Oh my god I was crying because I was anxious. I was scared for my life.”*

One potential explanation for the different experiences of pain may be that participants who had slighter quicker infusions (range 9-15 minutes) reported significantly less pain than participants whose infusions took slightly longer (range 16-25 minutes).

### Experiences and wellbeing in the days and weeks following SCIP

The Research Nurse observed that over half of participants had some discomfort one day following dosing and some participants had redness or bruising. However, by day two participants reported less pain, mean NRS 1.7 (range 0-5), and by day three all participants were free of discomfort or pain and life had resumed back to normal. On day 28 post-dosing when participants reflected on their experience almost all reported that the pain on day one and two had not stopped them doing their usual activities. Matisse found that the infusion didn’t stop her doing anything:

> *“Only a little pain and bruising did not stop me from doing anything.”*

### Intramuscular BPG is more painful than SCIP

When compared to their normal IM injection most participants found SCIP significantly less painful, with provided pain scores for their last IM averaging NRS 5 (range 0-9). Following IM injections, several participants noted that the pain post-IM lasted several days and made it uncomfortable to sit down, sleep or participate in normal activities. Talia and Hana describe how they felt after their usual IM injection:

> *“With my injection I feel pain which lasts 3-4 days sitting down and sleeping. With the infusion one I didn’t have that pain”*
>
> *“Pain [from monthly injection] is like a sore muscle like I have been hit with a tennis ball. It is every time I put pressure on it like sitting down and this lasts for two to three days.”*

Mele and Matisse also found SCIP less painful than their normal IM injection:

> *“I prefer it on my tummy, as it took just takes two days for pain to go away but with the injection in my butt it takes a week as it gets sore every time I sit down”*
>
> *“This one is better [than normal] it does not hurt. My normal injection would be a 5” [Inserting the SCIP needle was rated 1]*

### Overwhelming yes to remain on three-monthly SCIP rather than IM BPG

Participants were asked on day 28 (day of regular IM BPG) and 70 (day returning to IM BPG) whether they would prefer SCIP or IM BPG. By day 70 all participants (excluding one) said they would prefer to have SCIP. The nurse noted a sense of real happiness from the participants at not having to go for regular IM BPG injection on day 28 and a palpable sense of relief in their expressions. The key reasons participants gave related to the convenience of having SCIP every three months and reduced pain. Nina and Mia were happy to remain on SCIP:

> *“I am super relieved, less admin, one less thing for me to think about.”*
>
> *“I’d prefer the infusion because I don’t have to get injections every month.”*

The one participant who preferred not to continue SCIP was a child who tolerated SCIP well but had such a good relationship with the nurse who delivered their regular IM BPG injection that they preferred to see her regularly.

## Discussion

This study described patient perspectives alongside nurse and researcher observations of a Phase-II trial investigating the delivery of SCIP for those currently receiving standard IM BPG treatment. Our results demonstrate an overwhelming preference by participants with ARF presently prescribed four-weekly secondary prophylaxis to receive less frequent delivery of penicillin via a SC infusion. Of 20 participants, 19 wished to remain on SCIP rather than return to their regular four-weekly injection. The young participant who preferred to remain on IM BPG tolerated SCIP very well but having formed a good connection with their usual nurse, preferred to continue seeing them regularly.

The high level of support for SCIP was due to lower pain levels experienced during the infusion and while some participants experienced discomfort on days one and two following dosing, the pain reported was less severe and of shorter duration than that of their usual injection. These findings reinforce findings from a randomised cross-over trial undertaken by Kado et al., [18] which reported a significantly higher median pain score for those receiving IM BPG (1 [0.25–2]) as opposed to SCIP (0.5 [0–1]), p=0.03) 48 hours following dosing [26]. In addition to preferring SCIP because of the reduction in pain, participants responded favourably to the longer duration (70-91 days versus 28 days) between dosing. This meant participants did not need to take as much time off work or school and participants felt enabled to do things like go on holiday.

By delivering BPG through a series of slow pushes there was a greater control, in comparison to the spring infuser used in SCIP-I, over the rate the BPG was delivered. This potentially led to slightly shorter infusion times; SCIP-I - mean infusion time of 22 minutes, range 16-29 minutes [20], compared to a mean of 15 minutes, range of 9-25 minutes, in SCIP-II. In addition, participants in SCIP-II reported slightly lower pain scores during the infusion in comparison to SCIP-I (median NRS 1.0, range 0-7 median compared to median NRS 2.5, range 0-8) [19]. Further improvements to the delivery of SCIP may be achieved by ensuring patients develop a rapport with SCIP providers to help reduce participant anxiety, to improve patient experience and potentially reduce pain. Facilitating a shorter infusion time, which was typically associated with lower pain and would offer greater convenience for patients and healthcare providers. This could be further supported by penicillin reformulations with a reduced dose volume and/or lower force of administration.

One limitation of the study is that we did not ask about pain tolerance. However, we did compare, and contrast pain experienced during and following SCIP to participants’ usual IM BPG (although the latter was reported in retrospect, as a recollection of the pain following their most recent IM BPG injection, which in most cases was four-weeks previously). Another limitation was that some participants were difficult to get feedback from while others found it hard to express how they were feeling. This may have been related to a whānau (family) member being present for support, potentially limiting what participants were willing to say. However, we experienced that whānau were very supportive in terms of helping the participants provide examples of experiences particularly for those participants who found it hard to share their experiences.

A strength of this study was the high level of engagement from participants, with 100% adherence. This was reflective of trust, communication and rapport developed between the study team, health practitioners, and participants. The open-ended nature of the interview questions meant participants were able to discuss challenges with regular IM BPG and day-to-day life. In addition, engagement of Kaupapa Māori Research principles allowed participants to understand the benefits of participation for themselves and others with ARF.

SCIP is preferred over IM BPG as a tolerable and acceptable route of administration for children and young adults with ARF in Wellington, NZ. Extending this study to support widespread use of SCIP, throughout NZ in patients requiring secondary prophylaxis over a prolonged period will enable better assessment of key measures. It would be useful to undertake an economic evaluation of SCIP in comparison to IM BPG. Current evidence suggests that SCIP should become the standard of care for most patients needing long-term prophylaxis. SCIP has the potential to improve adherence, prevent disease progression and death, transforming prophylaxis of ARF/RHD both in NZ and globally.

## Data Availability

Audio-recordings of interviews available on request

## Acknowledgements

We would like to thank all the participants and their whānau (families) for sharing their time and experiences to make this study possible. We would also like to thank the outpatient department at Kenepuru Hospital for sharing their space with us and the wonderful community nurses for supporting the study.

## Author contributions

JC; data collection, data analysis, data interpretation, writing original draft and editing. SE; data analysis, data interpretation, writing original draft and editing. HD; study funding and clinical oversight; EA; clinical oversight and data interpretation, JK, RB, SS; study conceptualization and design, funding. DM; clinical oversight, data collection. MB, JC; funding and data interpretation. LM, AA; study conceptualization and design, funding, data analysis, data interpretation. JB; study conceptualization and design, funding, data collection, data analysis, data interpretation, writing original draft and editing. All authors have read and reviewed the manuscript.

## Data sharing

Audio-recordings of interviews available on request.

